# Collaterals influence oxygen metabolism on admission MRI in acute ischemic stroke

**DOI:** 10.1101/2024.12.12.24318960

**Authors:** Paul Patural, Alexandre Bani-Sadr, Roberto Riva, Carole Frindel, Charles de Bourguignon, Marc Hermier, Delphine Gamondes, Emilien Jupin-Delevaux, Laurent Derex, Tae-Hee Cho, Laura Mechtouff, Norbert Nighoghossian, Yves Berthezene

## Abstract

**Introduction:** This study aimed to correlate cerebral collateral status and MRI-derived oxygen metabolism at admission in acute ischemic stroke (AIS) patients treated with mechanical thrombectomy.

**Methods:** The HIBISCUS-STROKE cohort (CoHort of Patients to Identify Biological an Imaging markerS of CardiovascUlar Outcomes in Stroke; NCT: 03149705), a single-center observational study, enrolled AIS patients for thrombectomy following MRI triage due to anterior circulation large vessel occlusion treated with mechanical thrombectomy. Dynamic-Susceptibility Contrast perfusion (DSC perfusion), Diffusion-Weighted Imaging (DWI) and penumbral volume (Tmax ≥ 6 secs) were post-processed to generate oxygen extraction fraction (OEF) and cerebral metabolic rate of oxygen (CMRO2) maps within DWI and penumbral anomalies, compared to contralateral areas. Collateral status, assessed via pretreatment digital subtraction angiography, was categorized as poor (0–2) or good (3–4) based on ASITN/SIR collateral grading system score from Higashida.

**Results:** Between October 2016 and October 2022, 321 participants were enrolled in the cohort. Of these, 184 (57.3%) were excluded due to missing admission DSC perfusion MRI in 61 patients and artifacts precluding collateral status evaluation on DSA. A total of 137 patients were included (mean age 71 years; 56.2% male). The median National Institutes of Health Stroke Scale (NIHSS) score was 15 (interquartile range [IQR]: 8.0–18.0), and the median time from symptom onset to admission was 96.0 minutes ([IQR]: 78.0–137.0). Patients with good collaterals (78) exhibited a smaller baseline infarct core (median 9.83 mL; (P<0.0001) less impairment cerebral blood flow (CBF) within the DWI lesion (median -65.89%; (P<0.0001), and less severe decrease CMRO2 within the ischemic penumbra (median -17.29%; (P=0.03). Good collaterals were independently associated with a smaller baseline infarct core volume and less decrease in CMRO2 within the ischemic penumbra (respectively OR = 0.94; 95% CI: [0.92; 0.96]; (P<0.0001), and (OR = 1.30; 95% CI: [1.06; 1.82]; P=0.001).

**Conclusion:** Good collaterals are associated with a smaller infarct core and better CMRO2 within the ischemic penumbra.

## Introduction

The main role of the collateral circulation is to maintain blood flow to the tissue that is normally supplied by an occluded vessel. This slows down the progression of tissue from penumbra to infarct, thus increasing the therapeutic time window, decreasing the final stroke volume, and decreasing the risk of haemorrhagic transformation. The rate of progression from penumbra to infarct greatly depends on the extent of the collateral circulation and ultimately determines whether the patient will be a fast or a slow progressor tissue. The persistence of blood flow to the blood clot also enhances the delivery of endogenous or exogenous tissue plasminogen activator, which increases the recanalization rate and decreases the re-occlusion rate ^1–4^. In addition, collaterals status supply may likely promote effective recanalization after EVT.

The cerebral metabolic rate of oxygen (CMRO_2_) is a critical factor of the ischemic tissue viability ^5^. CMRO_2_ levels can be influenced by the importance of collateral circulation in both the core ischemic and penumbral regions. Positron emission tomography (PET) has traditionally identified thresholds for CMRO_2_ in the penumbra and ischemic core ^5–7^. However, this examination is not feasible in the context of acute stroke thrombectomy.

Advancements in the post-processing techniques of dynamic-susceptibility contrast (DSC) perfusion MRI, commonly integrated into AIS protocols, now allow for estimating oxygen metabolism ^8^.

This study aimed to determine the relationships between collaterals and oxygen metabolic changes occurring in AIS patients eligible for EVT. We hypothesize that oxygen metabolism could be less affected in patients with good collaterals.

## Methods

### Data Availability Statement

The data supporting the findings of this study are available from the corresponding author upon reasonable request.

### Ethics Statement

This study was approved by the local ethics committee (Institutional Review Board No: 00009118). All participants or their legal guardians provided written informed consent, adhering to institutional guidelines.

### Study Population

The HIBISCUS-STROKE cohort (CoHort of Patients to Identify Biological an Imaging markerS of CardiovascUlar Outcomes in Stroke; NCT: 03149705), was a single-center, observational study that prospectively enrolled patients with AIS due to anterior circulation large vessel occlusion. Participants underwent mechanical thrombectomy following initial MRI assessment. Intravenous thrombolysis was administered as per eligibility. Post-thrombectomy recanalization was assessed using a modified Thrombolysis in Cerebral Infarct (mTICI)^9^ score of ≥ 2B. A systematic brain CT scan was performed 24±6 hours post-procedure for hemorrhagic transformation assessment, using the European Cooperative Acute Stroke Study II classification^10–11^.

A follow-up MRI was further performed at day 6. Functional outcome was evaluated using the 3-months mRs during a face-to-face visit with a stroke neurologist.

### Neuroimaging DSA

Collateral status was evaluated via prethrombectomy digital subtraction arteriography (DSA) and categorized as poor (ASITN/SIR collateral grading system scale, 0–2) or good (ASITN/SIR collateral grading system scale, 3–4) ^12^. ASITN/SIR collateral grading scale is detailed in **Supplementary Table 1**.

### Inclusion and Exclusion Criteria

Inclusion criteria were: 1) availability DSC perfusion MRI at admission and, 2) DSA of adequate quality for the evaluation for collateral status. Exclusion criteria were: 1) DSC perfusion MRI with artifacts and 2) incomplete DSA assessment without the capillary phase.

### MRI Data Acquisition

MRIs were performed using 1.5 or 3 Tesla Ingenia scanners (Philips Healthcare, Best, The Netherlands), including sequences such as DWI, T2-gradient echo, T2-FLAIR, 3D-time-of-flight angiography, and DSC perfusion MRI. For DSC perfusion, a standard dose of 0.1mmol/kg of contrast agent (Dotarem, Guerbet, France) was administered at 4 mL/s, followed by a 20 mL saline flush.

### Assessment of Volume Changes of Ischemic Lesions

Ischemic lesions were quantified using a semi-automated method (3D Slicer, https://www.slicer.org/) by an experienced investigator blinded to clinical data. DWI abnormalities were segmented using region-of-interest thresholding (apparent diffusion coefficient [ADC] < 620×10−6 mm²/s), with manual adjustments as needed ^10^. DSC perfusion MRI was post-processed using OleaSphere® (Olea Medical, France) to generate time-to-maximum (Tmax) maps, computed via delay-insensitive circular singular value deconvolution with automatic arterial input function selection. Tmax maps were co-registered to ADC maps using a non-linear, voxel-based approach with advanced neuroimaging tools ^11^. Penumbra masks were segmented based on voxels with Tmax ≥6s excluding those with ADC ≤620×10−6 mm²/s. Day 6 assessments involved non-linear co-registration of T2-FLAIR to admission DWI using advanced normalization tools.

### Generation of oxygen metabolism maps

Cerebral blood volume (CBV), cerebral blood flow (CBF), CMRO_2_ and oxygen extraction fraction (OEF) maps were derived from DSC-MRI using Cercare Medical Neurosuite software (Cercare Medical, Denmark). These maps were co-registered with ADC images to extract their values the infarct core, the ischemic penumbra, and corresponding contralateral regions using mirror masks. Variations in CBF, CMRO_2_, and OEF were quantified as percentage changes from the unaffected hemisphere, providing a relative assessment of metabolic disruption. Additional methodological details regarding map computation are available in the supplementary material.

### Statistical Analysis

Continuous variables are presented as medians with interquartile ranges (IQR), and categorical variables as percentages. The Mann-Whitney U test, Chi-square test or Fisher’s exact test, and Kruskal-Wallis test were used as appropriate.

Univariable and multivariable logistic regressions were performed to assess factors associated with good collaterals. The multiple variable model included covariates from the univariable analysis with P-values below 0.1. Given the observed multicollinearity between CBF and CMRO_2_ (ρ=0.37, P<0.0001), CBF was omitted from the multiple variable model. Therefore, adjustments were made for the volume of the ischemic core, an occlusion of the internal carotid artery occlusion, relative change in CBF within the ischemic core, relative change in CMRO_2_ within the ischemic penumbra and tandem occlusion. A threshold of P<0.05 determined statistical significance.

## Results

### Study Population

Between October 2016 and October 2022, 321 participants were enrolled in the cohort. Of these, 184 (57.3%) were excluded including 61 patients without DSC perfusion MRI at admission and 123 with poor quality DSA. The study flow-chart is available in **Figure 1**. As detailed in **Supplementary Table 1**, excluded participants were more likely to have ICA termination occlusion (P<0.001) or tandem occlusion (P<0.001), but less likely to have M1 segment of the middle cerebral artery occlusion (P=0.002). No other significant differences were observed compared to included participants.

**Figure 1.**
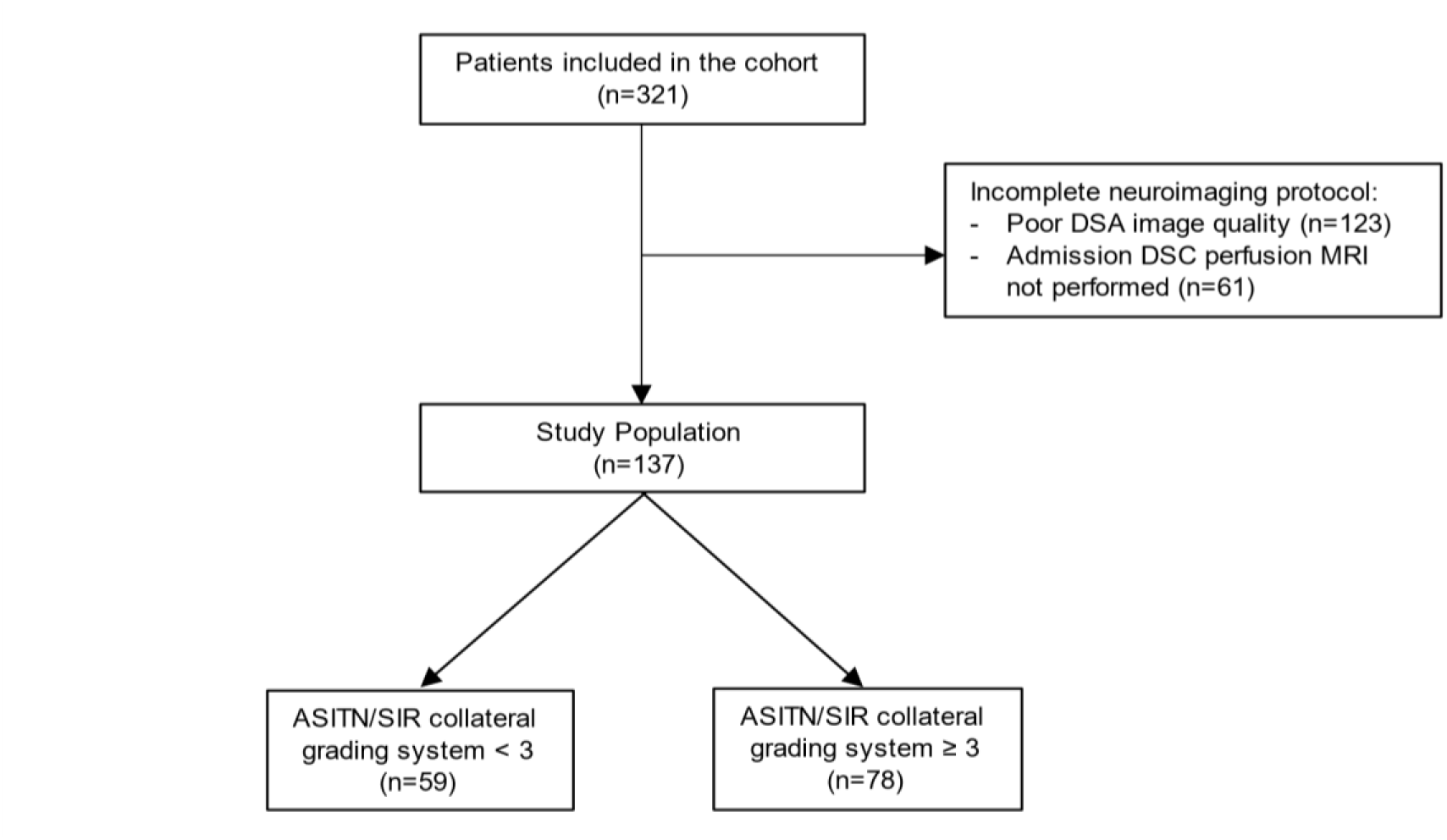
Flow-Chart of study population. <u>Abbreviations:</u> ASITN/SIR: American Society of Interventional and Therapeutic Neuroradiology/Society of Interventional Radiology; DSA: Digital Subtraction Arteriography; DSC: Dynamic-Susceptibility Contrast.

The study population consisted of the remaining 137 subjects (42.7%), with a median age of 71 years (IQR: [55.0, 82.0]), of whom 77 (56.2%) were males. The median National Institute Health Stroke Scale (NIHSS) score at admission was 15.0 (IQR: [8.0, 18.0]), and the median time interval from symptom onset to admission MRI was 96.0 minutes (IQR: [78.0, 137.0]).

### Descriptive Analyses

Among the 137 subjects, 78 patients (57.00%) had good collaterals (grade 3-4), while 59 patients (43.10%) had poor collaterals (grade 0-2). **Table 1** presents clinical and neuroimaging characteristics and outcomes according to cerebral collateral status.

**Table 1.**
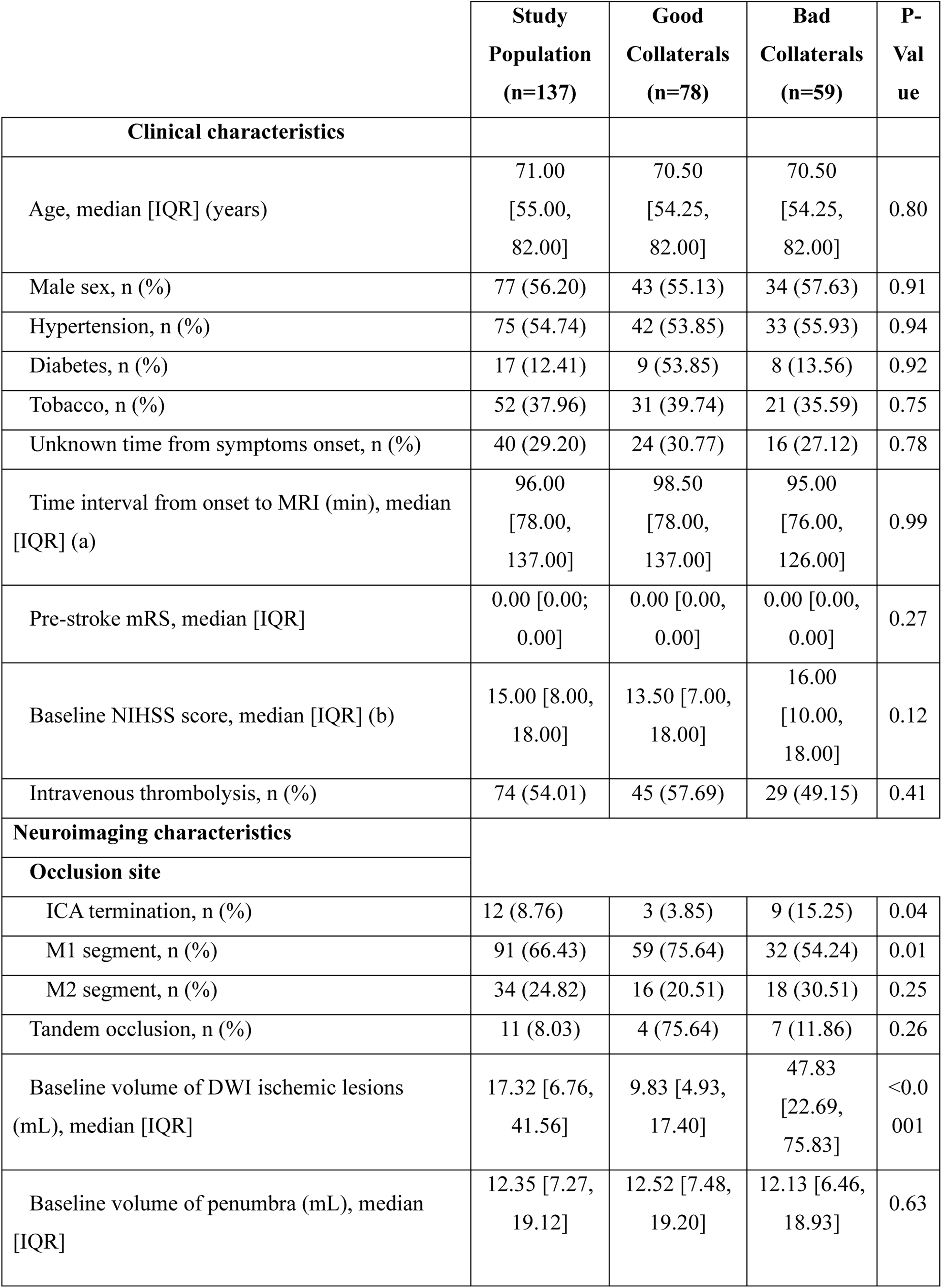
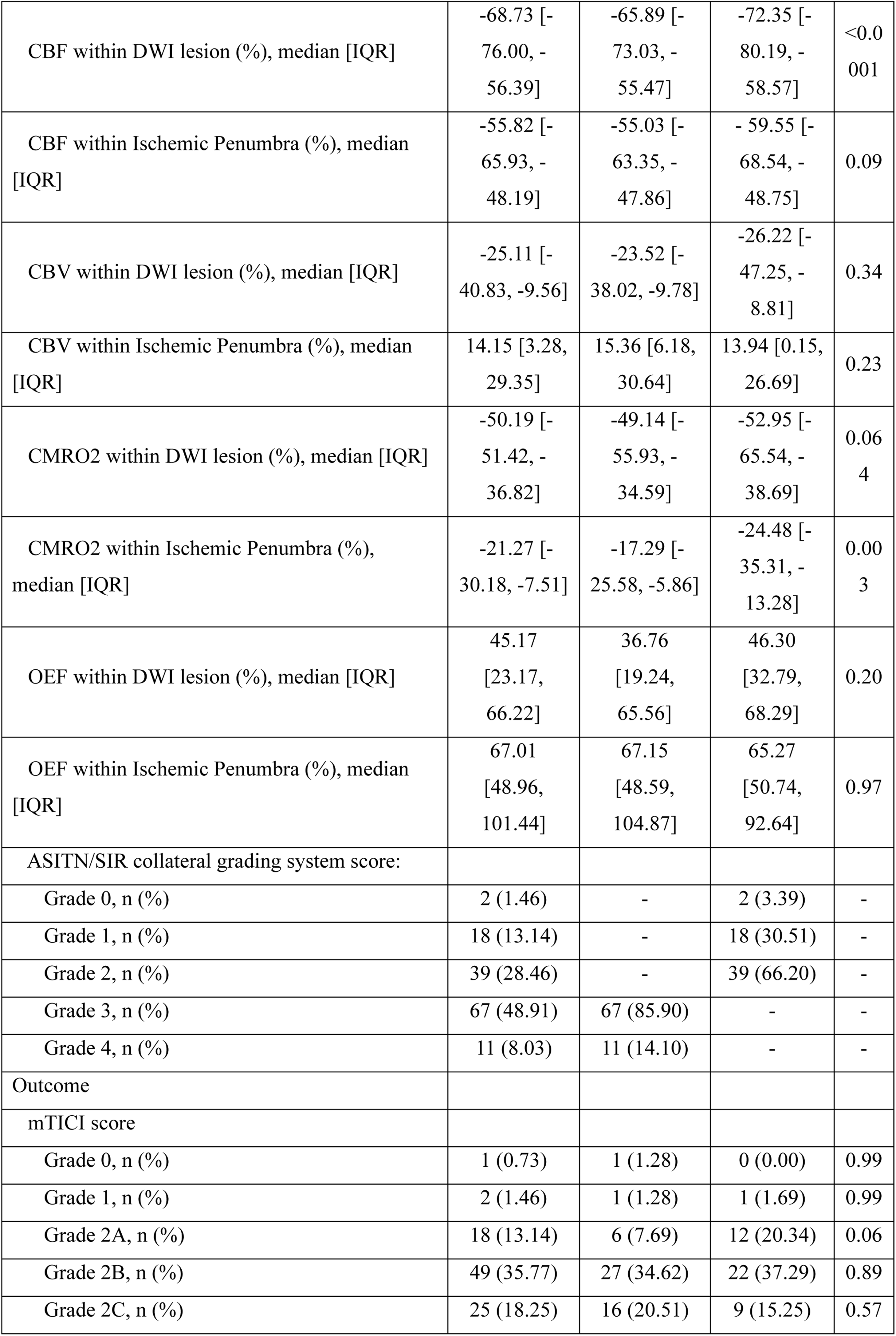
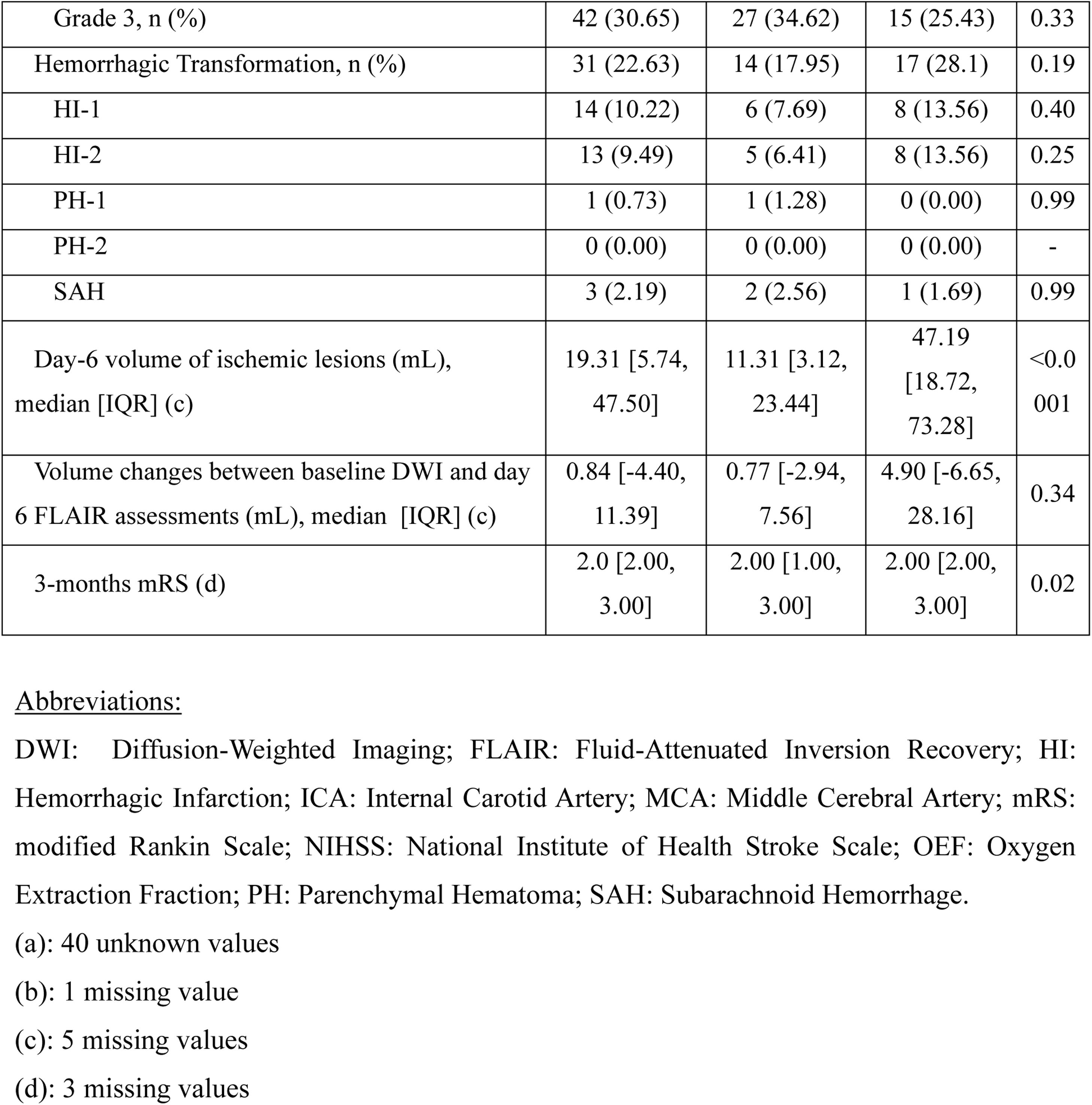
Descriptive analyses of study population according to the status of cerebral collaterals.

In the whole population, both CBF and CMRO_2_ decreased in DWI lesion (respectively median = -68.73 % IQR: [-76.00;-56.39] and -50.19 % IQR: [-51.42;-36.82]) and in ischemic penumbra (median = -55.82 %, IQR: [-65.93;-48.19], and -21.27 %, IQR: [-30.18;-7.51]), and OEF increased in DWI lesion (median = + 45.17, IQR: [+23.17;+66.22]) and increased in ischemic penumbra (median = +67.01 %, IQR: [+48.96;+101.44]).

In comparison to patients with poor collaterals, those with good collaterals exhibited a smaller infarct core (median 9.8 mL, IQR: [4.9, 17.4] vs. 47.8 mL, IQR: [22.7, 75.8]) (P<0.0001) and a smaller final infarct volume at day 6 (median: 11.3 mL, IQR: [3.1; 23.4] vs. 47.2 mL, IQR: [18.7; 73.3]) (P<0.0001). Baseline penumbral volume showed no significant difference (median: 12.52 mL, IQR: [7.48; 19.20] vs. 12.13 mL, IQR: [6.46; 18.93], P= 0.63).

Compared to patients with poor collaterals, those with good collaterals demonstrated less severe decrease in CMRO_2_ within the ischemic penumbra (median -17.3%, IQR: [-25.6, -5.9] vs. -24.5%, IQR: [-35.3, -13.3]) (P=0.03). A non-significant trend was observed within the ischemic core (median: -49.1%, IQR: [-55.9, -34.6] vs. -52.9%, IQR: [-65.5, -38.7]) (P=0.06).

Patients with good collaterals exhibited less severe impairment in CBF within the ischemic core (median -65.9%, IQR: [-73.3, -55.5] vs. -72.4%, IQR: [-80.2, -58.6], P<0.0001). They also tended to have less severe impairment within the ischemic penumbra (median -55.03 %, IQR: [-63.35, -47.86] vs. -59.55%, IQR: [-68.54, -48.75], P=0.09).

OEF changes did not differ significantly according to collaterals status within ischemic core (median 36.76%, IQR: [19.24, 65.56] vs. 46.30%, IQR: [32.79, 68.29] (P=0.20) and ischemic penumbra median: 67.15%, IQR: [48.59, 104.87] vs. 65.27%, IQR: [50.74, 92.64] (P=0.97)).

An illustration of the metabolic abnormalities according to collateral status is presented in **Figure 2**.

**Figure 2:**
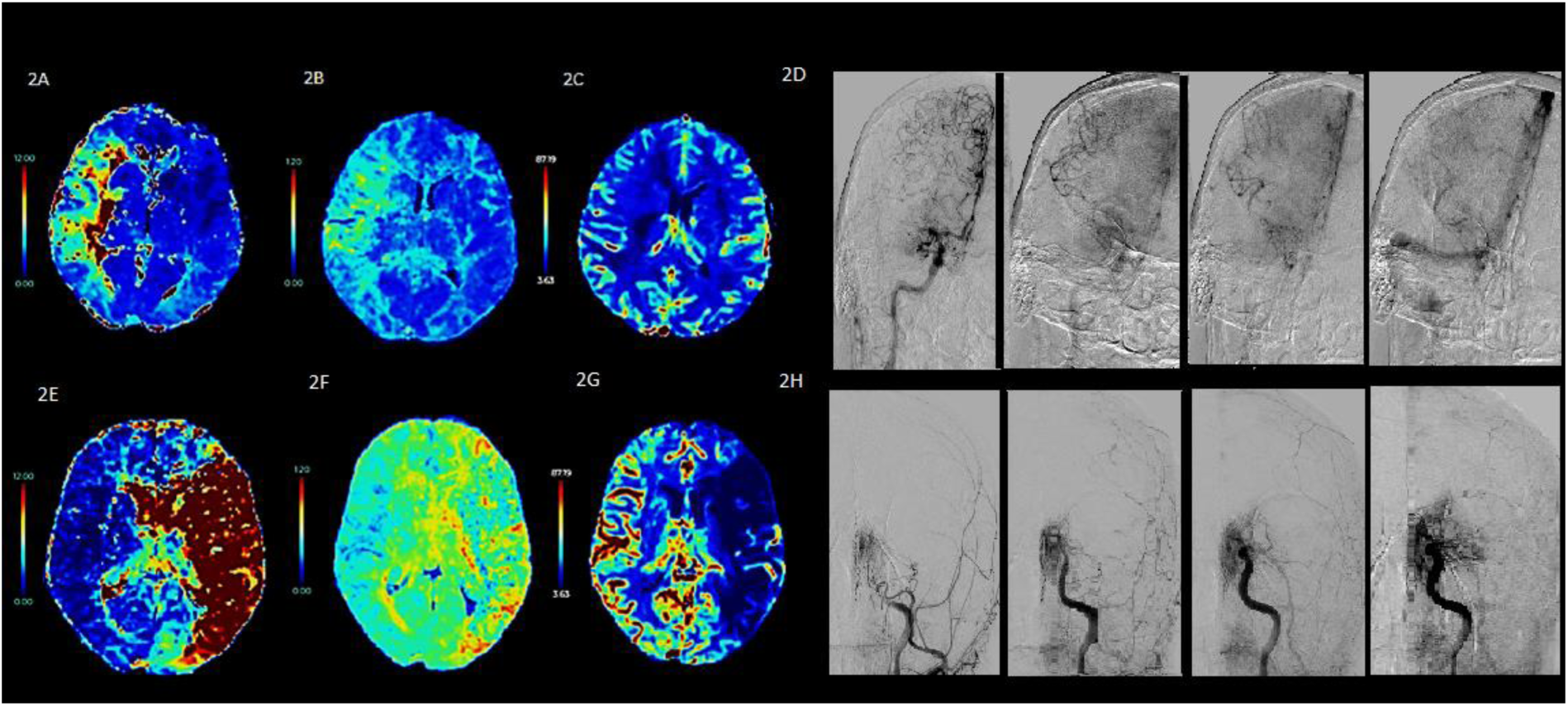
illustration of the metabolic abnormalities encountered in patients after stroke, according to collateral status. 2A-2D: MRI abnormalities in a patient with good collaterals and its DSA: Tmax (2A), OEF (2B), CMRO_2_ (2C) and DSA (2D). 2E-2H: MRI abnormalities in a patient with bad collaterals and its DSA: Tmax (2E), OEF (2F), CMRO_2_ (2G) and DSA (2H). In patients with good collaterals, a less pronounced decrease in cerebral metabolic rate of oxygen was noted within the ischemic penumbra. <u>Abbreviations:</u> CMRO_2_: Cerebral Metabolic Rate of Oxygen; DSA: Digital Subtraction Angiography; OEF: Oxygen Extraction Fraction; Tmax: Time-To-Maximum.

### Factors Associated with Cerebral Collateral Status

At univariate logistic regressions, good collaterals were associated with smaller ischemic cores (odds ratio (OR) = 0.94, 95% CI: [0.92; 0.96], P<0.0001), less severe decrease in CBF (OR = 1.26, 95% CI: [1.03; 1.62], P=0.04) and CMRO_2_ (OR = 1.44; 95% CI: [1.15; 1.85]; P=0.002).

At multiple variable analysis, good collaterals remained associated with smaller ischemic cores (OR = 0.94; 95% CI: [0.92; 0.96]; P<0.0001) and less severe impairment of CMRO_2_ within the ischemic penumbra (OR = 1.30; 95% CI: [1.06; 1.82]; P=0.001). **Table 2** presents the unadjusted and adjusted OR.

**Table 2.**
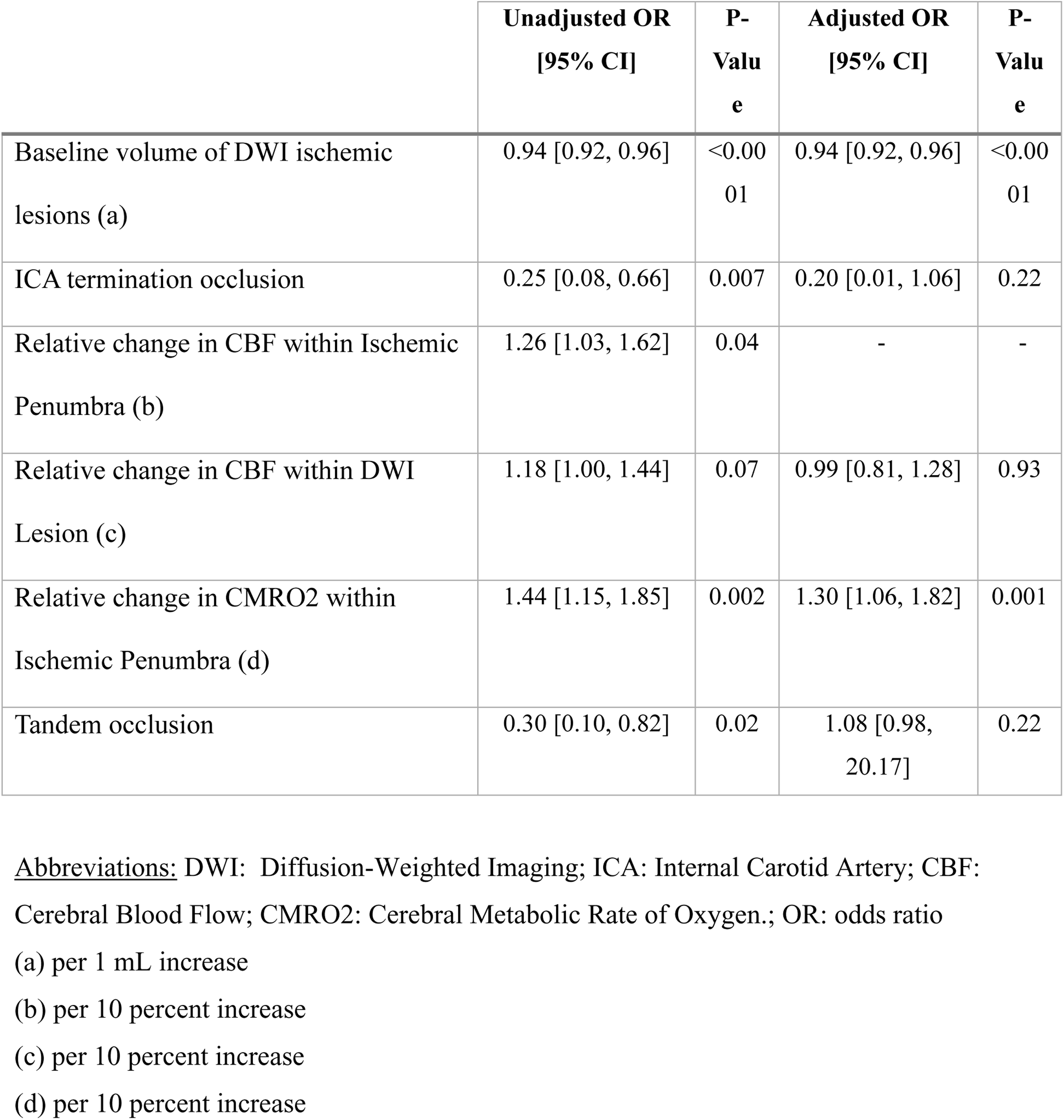
Multiple variable logistic regression of factors associated with good collaterals.

## Discussion

In this study, we observed that good collaterals were independently associated with smaller ischemic cores and less severe impairment of cerebral metabolic rate of CMRO_2_ within the ischemic penumbra. In early PET studies, the penumbra has been defined as an area with relatively preserved CMRO_2_ in contrast to the ischemic core. In agreement, we found a significant decrease in CMRO_2_ in the infarct core and a moderate decrease in the penumbra. Moreover, still in agreement with PET, we found an increase of OEF within penumbra.

Evaluation of the penumbra using CMRO_2_ in AIS is relatively understudied on MRI. Oxygen metabolism maps derived from DSC perfusion MRI can be used in acute clinical settings. Unlike other methods such as QSM-OEF, they do not require additional MRI sequences and do not extend the post-processing duration.

Authors have suggested CMRO_2_ thresholds using MRI in animals to distinguish ischemic lesions from penumbra, similar to findings in previous PET studies ^13^.

In this study, we observed that good collateral circulation was independently associated with smaller ischemic core, although similarly to Nannoni et al, we found no correlation between collaterals and penumbra volumes following AIS ^14^. Most studies have predominantly focused on how collaterals sustain the penumbra after a stroke event ^15–20^.

The main result of this study is that there is a moderate decrease in CMRO_2_ in the penumbra (compared to the ischemic core) and that this decrease is less pronounced in the case of good collaterals.

The CMRO_2_ reflects the absolute value of the oxygen consumption of brain tissue^21^. It depends on CBF, OEF and arterial oxygen content. Collaterals, by attenuating the decrease in CMRO2, help sustain the ischemic penumbra. In other words, good collaterals appear to mitigate the decline in oxygen metabolism within the penumbra, thereby preserving the tissue’s ability to utilize oxygen more efficiently.

Seifert K and Heit JJ indicates that good collaterals are correlate with slower infarct evolution ^22^. Our study demonstrates the direct interplay between the collateral status and the state of the cerebral oxygen metabolism within the ischemic penumbra. Compared to patients with poor collaterals, those with good collaterals had less pronounced decrease in CMRO_2_ which is the primary determinant of the viability of the ischemic penumbra ^5; 20^.

Interestingly, OEF within the penumbra did not differ significantly between cases with good and poor collaterals.

Previous studies have shown that OEF increases in both the ischemic lesion area and the penumbra^23–25^. However, OEF initially experiences a lesser increase in the ischemic core compared to the penumbra, suggesting a complex interplay between oxygen demand and supply in the core ischemic area. Nevertheless, as the ischemic event progresses, OEF subsequently declines in both the core and penumbral regions, indicating a dynamic adjustment in oxygen extraction efficiency.

Despite the rise in OEF in the peri-infarct region, OEF alone was found to be a relatively weak predictor of tissue outcome ^27^. In contrast, CMRO2 has been demonstrated to be more effective in distinguishing tissue viability after AIS using PET ^26–29^.

This study has several limitations beyond its single-center nature and the retrospective assessment of oxygen metabolism maps. The primary constraint arises from the absence of validation of DSC-MRI-derived oxygen metabolism maps with PET. However, direct comparisons of small sample sizes suggest a strong correlation, and the methodology has proven robust against variations in signal-to-noise ratio and arterial delay^30–31^. Additionally, OEF and CMRO2 derived from DSC perfusion MRI represents a relative rather than a quantitative assessment of OEF and CMRO_2_ values.

## Conclusion

In conclusion, we observed that good collateral circulation was independently associated with less severe impairment of cerebral metabolic rate of CMRO_2_

This highlights the neuroprotective role of collaterals in mitigating ischemic damage.

## Declarations

None

## Supplementary materials

**Supplementary Table 1:** Comparative analysis of the included and excluded populations.

|  | **Study**<br>**Population**<br>**(n=137)** | **Excluded**<br>**Population**<br>**(n=184)** | **P-**<br>**Value** |
| --- | --- | --- | --- |
| **Clinical characteristics** |  |  |  |
| Age, median [IQR] (years) | 71.0 [55.0, 82.0] | 71.0 [59.3; 80.8] | 0.97 |
| Male sex, n (%) | 77 (56.2) | 98 (53.9) | 0.76 |
| Unknown time from symptoms onset, n (%) | 40 (29.2) | 48 (26.4) | 0.80 |
| Time interval from onset to MRI (min), median [IQR] (a) | 96.0 [78.0, 137.0] | 108.0 [80.0, 165.0] | 0.23 |
| Baseline NIHSS score, median [IQR] (b) | 15.0 [8.0, 18.0] | 14.0 [8.0; 19.0] |  |
| Intravenous thrombolysis, n (%) | 74 (54.0) | 102 (56.0) | 0.67 |
| **Neuroimaging characteristics** |  |  |  |
| Occlusion site |  |  |  |
| ICA termination, n (%) | 12 (8.8) | 61 (33.5) | <0.001 |
| M1 segment, n (%) | 91 (66.4) | 88 (48.4) | 0.002 |
| M2 segment, n (%) | 34 (24.8) | 33 (18.1) | 0.19 |
| Tandem occlusion, n (%) | 11 (8.0) | 41 (22.5) | <0.001 |
| Baseline volume of DWI ischemic lesions (mL), median [IQR] (c) | 17.3 [6.8, 41.6] | 12.9 [4.6; 32.0] | 0.08 |

(a): 88 unknown values
(b): 1 missing value
(c): 24 missing values
Abbreviations: DWI: Diffusion-Weighted Imaging; ICA: Internal Carotid Artery; IQR: Interquartile Range; MCA: Middle Cerebral Artery; NIHSS: National Institute of Health Stroke Scale.

## Dynamic-Susceptibility Contrast MRI post-processing

The dynamic-susceptibility MRI were post-processed automatically using the Cercare Medical Neurosuite software (Cercare Medical, Denmark, version release-2023-02-01-03). In short, the processing pipeline consisted of image reading and sorting, followed by slice time correction through standard spline methodologies, and motion correction via the SimpleElastix software^1^ (SimpleElastix, version 1.1.0) using affine registration to the initial 3D volume of the perfusion series. After conversion of the intensity-time-curves to concentration-time-curves for each voxels, an arterial input function (AIF) was automatically found through a slightly updated version of the hierarchical K-means clustering method described in Mouridsen et al^2^.

With the availability of the AIF and voxel-wise concentration-time-curves, the computation of hemo-dynamic parameters was performed using a so-called parametric or vascular model^3–5^ deconvolution method. With this method, voxel-wise estimates of flow and the so-called residue function are computed from the general indicator-dilution theory-based equation^6^ :

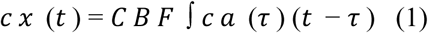

Where *c t* (*t*) is the observed concentration in voxel x at time t, CBF is the cerebral blood flow, *c a* (*t*) is the AIF, and *R x* (*t* − *τ*) is an unobserved so-called residue function specific to voxel x measured at time t – t. The vascular model works from the principle that the residue function originates from a gamma-distribution of vascular transit times, with the residue function specifically given as

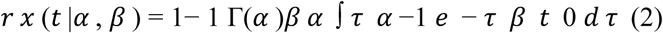

Where a and b are parameters and G(a) is the gamma function taken at the argument a. The transit time distribution is obtainable as the negative of the residue function derivative:

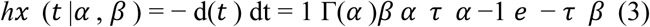

The model parameters, including CBF, a, b, and a delay measure are estimated voxel-wise via a so-called Bayesian method4. From the residue function, a series of hemo-dynamic parameters can be computed, including (i) the mean transit time (MTT), (ii) the so-called capillary transit time heterogeneity (CTH), (iii) a model-based maximum oxygen extraction fraction (OEF(model-based), or OEFmax), and (iv) a model-based relative maximum cerebral metabolic rate of oxygen (rCMRO2(model-based) or rCMRO2 max). For further information, please visit the papers by Jespersen and Østergaard5 and Mouridsen et al^4^.

All output images were available as DICOM image series for further analysis.

